# Increasing Firearm Deaths in The Youngest Americans: Ecologic Correlation with Firearm Prevalence

**DOI:** 10.1101/19009191

**Authors:** Archie Bleyer, Stuart Siegel, Charles R. Thomas

**Affiliations:** Knight Cancer Institute and Department of, Radiation Medicine, Oregon Health and Science University, Portland, Oregon; McGovern Medical School, University of Texas, Houston, Texas; Los Angeles, California

**Keywords:** firearms, firearm accidents, mortality, children, United States

## Abstract

**Background:** In the United States (U.S.), the overall death rate in 1-4 year-olds had been steadily declining until 2011, after which it ceased to improve. To understand this trend reversal, we investigated trends in the causes of their deaths.

**Methods:** Mortality data were obtained from the U.S. Centers for Disease Control and Institute for Health Metrics and Evaluation, firearm background check data from the National Instant Criminal Background Check System, and civilian firearm prevalence from the Small Arms Survey.

**Findings:** In 1-4 year-olds, the rate of fatal firearm accidents during 2002-2017 increased exponentially at an average rate of 6.0%/year (p=0.0003). The rate of increase was the greatest of all evaluable causes of death in the age group. Both the rate increase and most recent absolute rate in firearm accidental deaths in young children were correlated with the concurrent corresponding rate of firearm background checks (p = 0.0002 and 0.003, respectively). Also, the firearm accidental death rate in countries with high civilian firearm prevalence was correlated with the number of guns per civilian population (p=0.002).

**Interpretation:** Prior to 2004, the childhood firearm death rate did not increase during the Federal Assault Weapons Ban. Since 2004 when the Ban ended, the steadily increasing rate of sales and concomitant availability of, and access to, firearms in the U.S. has been associated with an increase in fatal firearm accidents in its youngest children. The acceleration of firearm deaths and injuries among young Americans requires urgent, definitive solutions that address firearm prevalence.

**Funding:** No external funding.

**KEY POINTS:** *Question:* In the U.S., how has the escalation of both firearm sales and firearm death rates affected the country’s youngest population?

*Findings:* While the steadily increasing rate of sales and concomitant availability of, and access to, firearms in the U.S. has increased since 2004, fatal firearm accidents in 1 to 4 year-olds increased exponentially and at a rate greater than all other evaluable causes of death in the age group.

*Interpretation:* The ominous acceleration of firearm deaths and injuries among young Americans requires urgent, definitive solutions from multiple stakeholders to effectively reduce firearm access.

## INTRODUCTION

According to the Institute for Health Metrics and Evaluation (IHME) Global Disease Burden (GBD) database,^1^ the United States (U.S.) has had a much slower reduction in the overall death rate in 1-4 year-olds since 1997 than all other countries of high socio-demographic index. By 2014 the rate was 43%-46% higher in the U.S. (Fig. 1**B**) and highly statistically significant (p < 0.0001) (Fig. 1**A**).^1^ As of 2017, only Brunei of the 39 other countries of high socio-demographic status had a higher rate.^1^ Even more concerning, the death rate among 1-4 year-olds in the U.S. ceased to decline since 2011 (Fig. 1**B**). Why has the death rate in the youngest Americans failed to improve while it has continued to do so in other countries?

**Figure 1.**
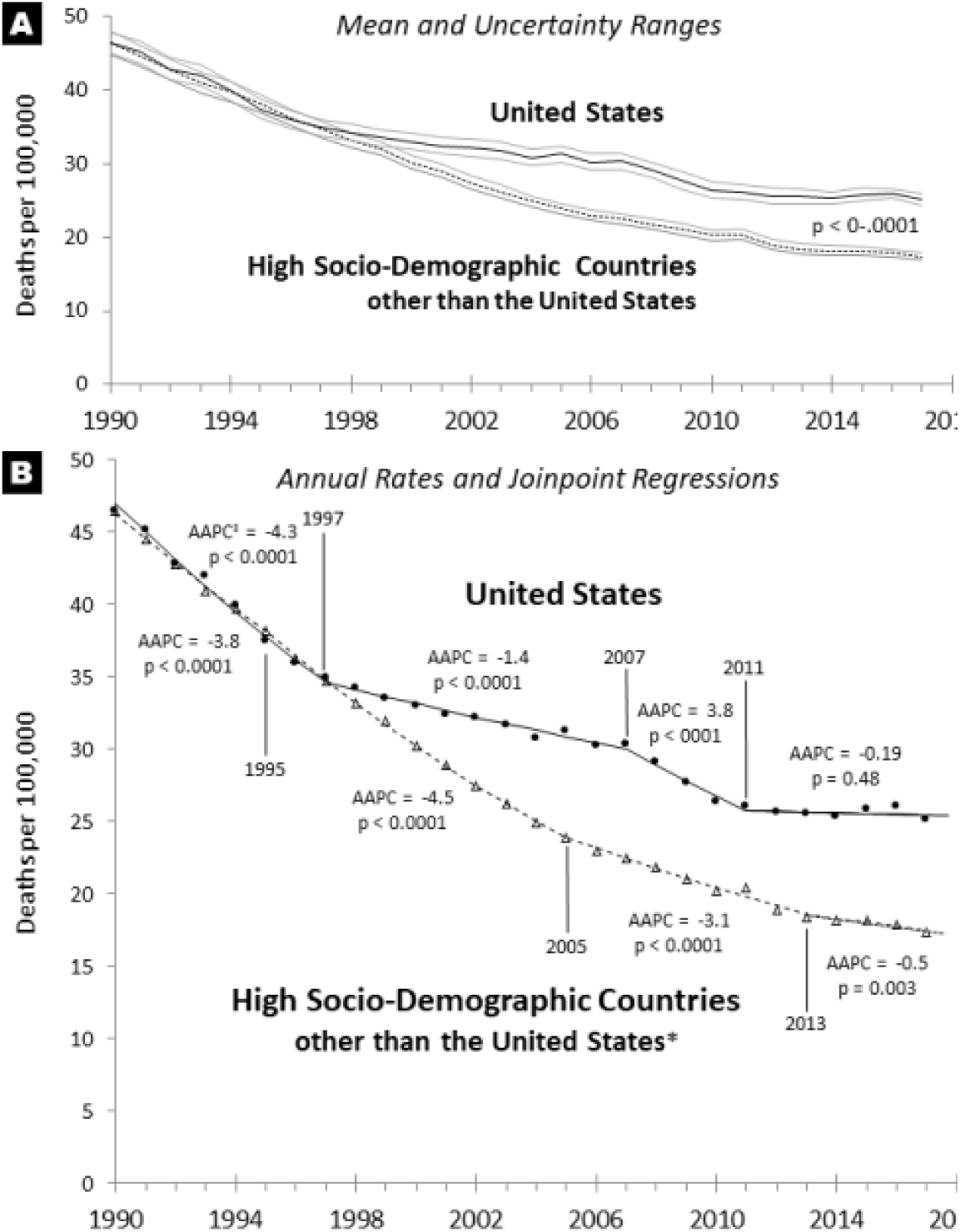
Comparison of the Annual Overall Death Rate in 1-4 Year-Olds during 1990-2017 in U.S and Countries of High Socio-Demographlc Index * Other Than the U.S. **A. Mean and Uncertainty Ranges** **B. Joinpolnt^9^/AAPC† Analysis** ‡ AAPC - average annual % change, based on constant variance (homoscedasticity) assump * 39 countries identified by IHME (http://ghdx.healthdata.Ofg/sites/default/files/record-attached-files/lHME_GBD_2017_S950_2017.zip): Andorra, Australia, Austria, Belgium, Brunei, Canada, Croatia, Cyprus, Cze Republic, Denmark, England, Estonia, Finland, France, Germany, Greece, Iceland, Ireland, Italy, Japan, Latvia, Lithuania, Luxembourg, Malta, Netherlands, New Zealand, Northern Ireland, Norway, Poland, Scotland, Singapore, Slovakia, Slovenia, South Korea, Spain, Sweden, Switzerland, Taiwan, Wales. <u>Data Source:</u> IHMF GRD^1^

Cunningham and colleagues reported that in 2016 firearms were the most common cause of death in urban U.S. children and teenagers.^2^ They reported the rate to be 37 times higher in the U.S. than in 12 other high-income countries and 5 times higher than seven low-to-middle-income countries. A recent study in Los Angeles County found that unintentional firearm injuries were more common in younger children, more frequently caused by a firearm from within the home, and more likely to involve friend/family.^3^ We therefore examined U.S. and global firearm mortality data and found that the firearm accident fatality rate increased exponentially since the early 2000s in children <5 years of age. We, therefore, tested the hypothesis that increasing firearm prevalence in the U.S. is a cause of the firearm accident mortality rate increase in young children.

## METHODS

### Death rates and causes

We obtained annual mortality data available since 1999 for the U.S. from the CDC Wide-ranging ONline Data for Epidemiologic Research (CDC WONDER)^4^ and for other countries from the IHME GBD.^1^ The CDC WISQAR database^4^ provided injury mortality data for the U.S. prior to 1999. The International Classification of Disease (ICD) 10 Codes^5^ for type of firearm death were W32-34 for unintentional; X93-95 for assault, Y22-24 for unspecified/undetermined, W32,X93,Y22 for handgun, W33,X94,Y23 for larger firearms, and W34,X95,Y24 for other/unspecified firearm.

### Firearm sales and civilian firearm prevalence

We estimated national firearm sales from firearm background checks since 1999 published by the National Instant Criminal Background Check System.^6^ Global data on estimated civilian gun prevalence were obtained from the Small Arms Survey.^7^

### Trend analysis

We analyzed time trends with the Joinpoint Regression Program^8^ that identifies when a trend changes to another trend, the probability range of the inflection, and the average annual percent change (AAPC) and p-value for each trend detected. We applied joinpoint analysis with weighted least squares option, logarithmic transformation, standard errors for annual death rates, and constant variance (homoscedasticity) for death rates without standard error values.

## RESULTS

### Firearm Death Rate Trends in 1 to 4 Year-Olds in the U.S. during 1999-2017

The national firearm death rate in American 1-4 year-olds fell progressively during 1999-2003, after which it increased exponentially (AAPC = 3.0, p = 0.003) (Fig. 2**A**). During 2003-2017, 60% of the increase in the firearm death rate in 1-4 year-olds was due to fatal accidents (unintentional causes), 27% to homicides, and 13% to undetermined and other causes (Fig. 2**A**). During 2002-2017 the firearm accidental death rate in 1-4 year-olds increased exponentially at an average AAPC of 6.0 (p = 0.0003) (Fig. 2**A**). Compared to the rate during 1999-2004 before the increase in the firearm background check rate, the firearm accidental death rate in 1-4 year-olds increased to 2.5 times higher in 2016-2017, according to the area under the death rate vs. age curves (Fig. 3**A**). For ages 2, 3 and 4, the 2002-2017 AAPC was 9.1, 11.0 (p < 0.05), and 8.5 (p < 0.05), respectively (Fig. 3**B**).

**Figure 2.**
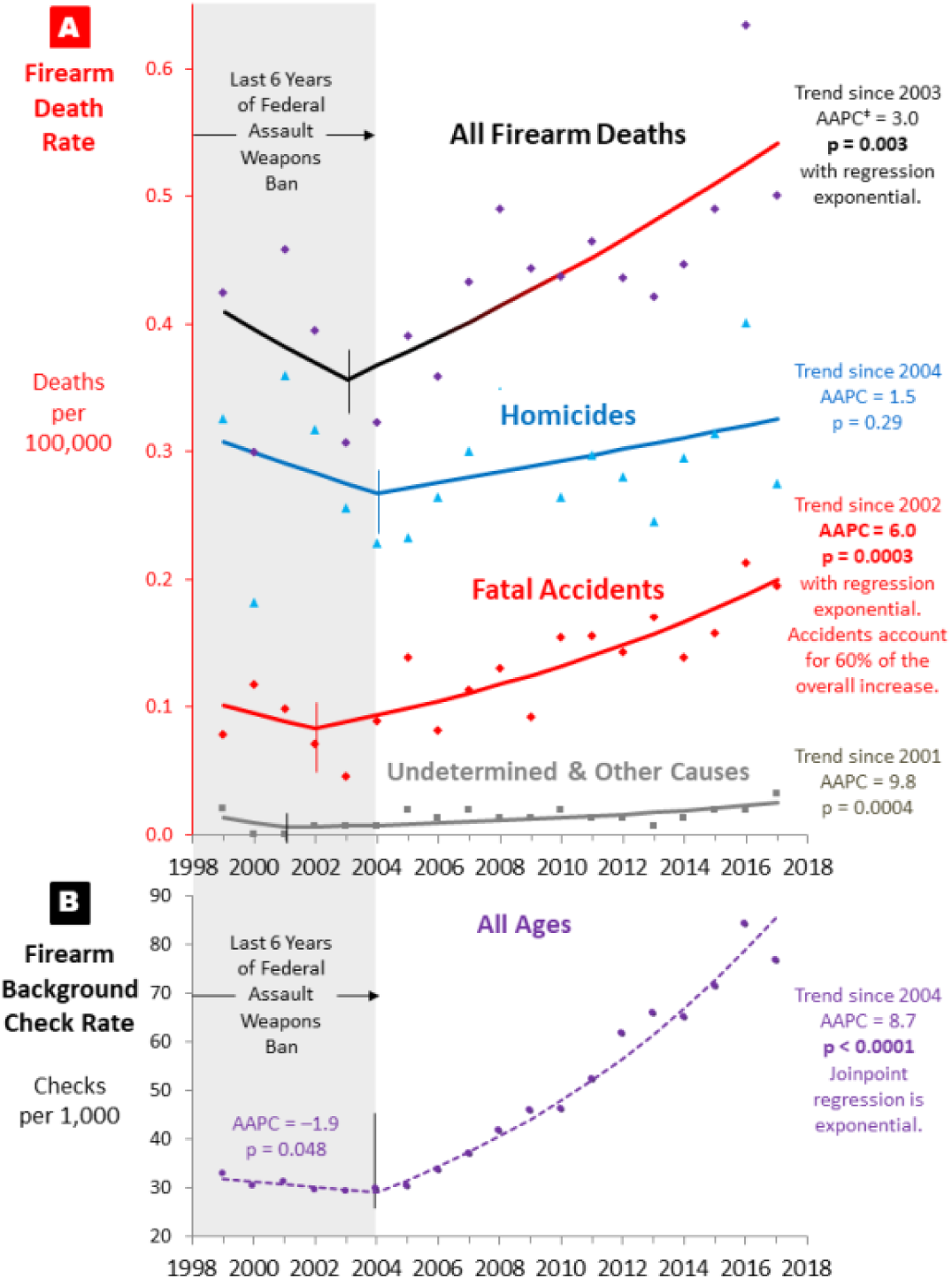
Joinpoint^9^/AAPC^‡^ Analysis of Annual Ratos during 1999-2017 in U.S. of: **A. Firearm Deaths in 1-4 Year-Olds, by Type of Firearm Death** **B. A Il-Age Firearm Background Check Rate** ^‡^ AAPC - Average Annual Percent Change Gray zone: Last 6 years of the 1994-2004 Federal Assault Weapons Ban <u>Data Source:</u> CDC WONDER,^4^ NICS^6^

**Figure 3.**
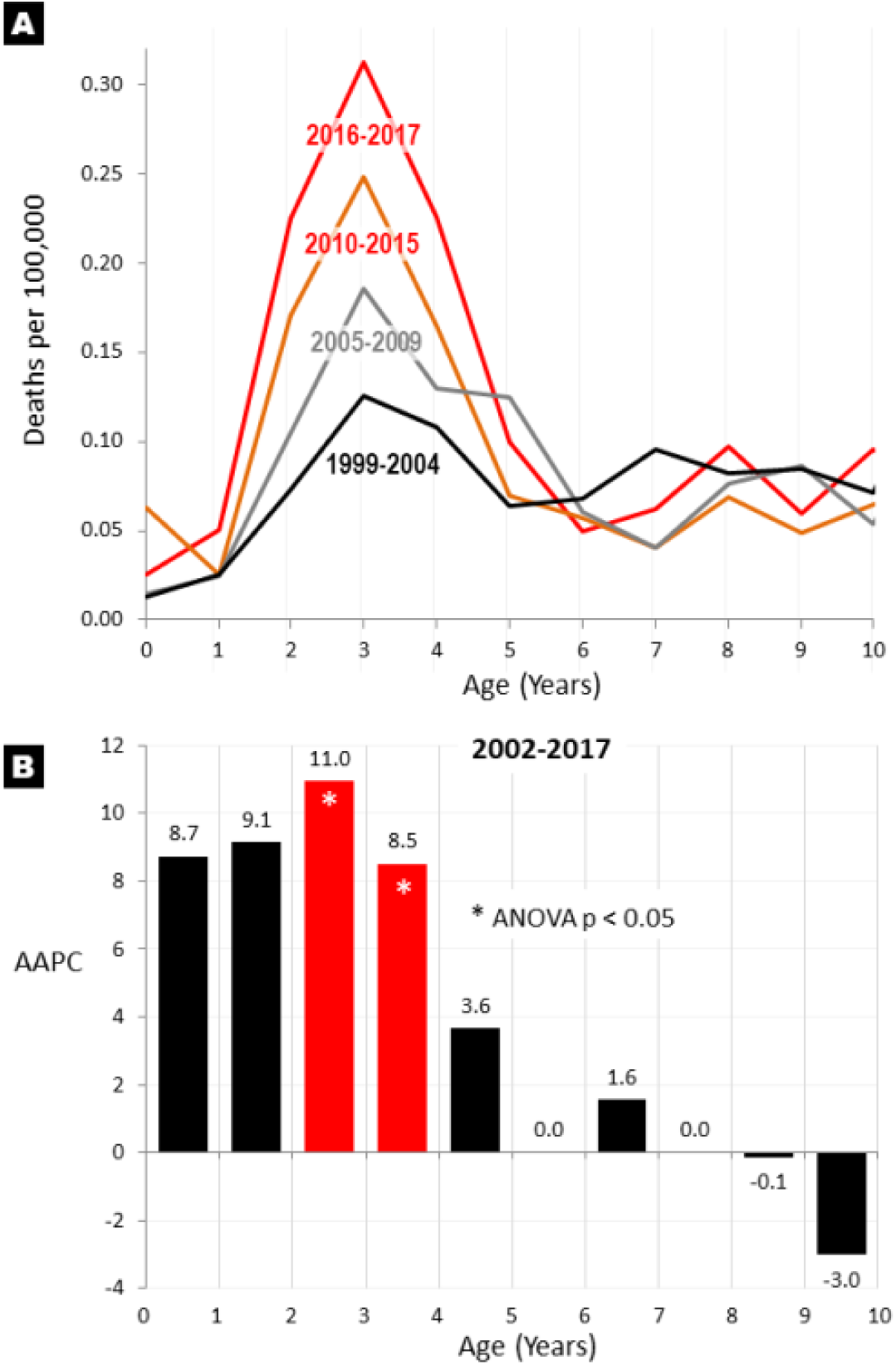
Firearm Accidental Death Rate by Single Year of Age from Birth to 10 Years, U.S. **A. By Era during 1999-2017.** **B. Average Annual Percent Change (AAPC) during 2002-2017.** <u>Data Source:</u> CDC WONDER^4^

Given the 2002-2017 era as having a continuously increasing accidental firearm death rate in 1-4 year-olds, this cause of death was compared in Table 1 with all of the leading causes of death in the age group during those years. For causes that had more than 50 deaths in the age group during 2002-2017, firearm accident fatalities had the greatest increase of the leading 43 evaluable causes of death (Table 1), 40% more than the cause with the 2^nd^ greatest increase. The 3^rd^ greatest increase was firearm deaths due to causes other than accidents, primarily undetermined causes and some homicides. It is likely that some of the undetermined causes were accidents that were not witnessed and thus not certain enough to be classified as an accident. Of the 43 leading evaluable causes, motor vehicle accidents in which 1-4 year-olds died as occupants had the greatest decrease (AAPC = − 8.14, p < 0.0001). Since 1999-2004 when 1-4 year-olds were >4 times likely to die in a motor vehicle than of a firearm, the former declined at an AAPC of −10.5 (p < 0.0001) such that by 2014-2017 1-4 year-olds died from bullets at the same rate as from motor vehicle accidents (Fig. 4). During 2016-2017, one in every 122 deaths that occurred in 1-4 year-olds occurred as a result of a firearm accident (Table 1).

**Table 1.** The 43 Leading Causes of Death in 1-4 Year-Olds during 2002-2017: Their Rate Trends and Proportion of All Deaths in 2016-2017

| Cause in trend rank order |  | AAPC* in Death Rate<br>during 2002-2017 | % of all Deaths<br>in 2016-2017 |
| --- | --- | --- | --- |
| 1 | **Firearm accident** | **7.46** | **0.8%** |
| 2 | Intestinal infections | 5.31 | 0.5% |
| 3 | **All other firearm** (primarily undetermined, some homicides) | **2.70** | **1.6%** |
| 4 | Other and unspecified infectious or parasitic diseases and their sequelae | 2.47 | 2.3% |
| 5 | Infections of kidney | 2.20 | 0.0% |
| 6 | Tuberculosis | 1.75 | 0.1% |
| 7 | Diabetes mellitus | 1.40 | 0.2% |
| 8 | Poisoning | 1.39 | 1.5% |
| 9 | Symptoms, signs, abnormal clinical or lab results not elsewhere classified | 0.74 | 6.8% |
| 10 | Other diseases of respiratory system | 0.52 | 2.5% |
| 11 | Machinery | 0.41 | 0.3% |
| 12 | Struck by or against | -0.08 | 0.7% |
| 13 | Hernia | -0.26 | 0.1% |
| 14 | Natural/Environmental | -0.38 | 0.8% |
| 15 | Cerebrovascular diseases, including stroke | -0.58 | 1.5% |
| 16 | Cut/Pierce | -0.62 | 0.4% |
| 17 | Other land transport | -0.90 | 0.4% |
| 18 | Other acute lower respiratory infections | -0.95 | 0.5% |
| 19 | Suffocation | -0.97 | 3.5% |
| 20 | Drowning | -1.15 | 11.2% |
| 21 | Unspecified Injury | -1.17 | 3.8% |
| 22 | Malignant neoplasms | -1.22 | 8.9% |
| 23 | Complications of medical and surgical care | -1.88 | 0.4% |
| 24 | Anemias | -1.89 | 0.6% |
| 25 | Chronic lower respiratory diseases | -1.97 | 1.0% |
| 26 | Congenital malformations, deformations and chromosomal abnormalities | -1.98 | 10.8% |
| 27 | Diseases of Heart | -2.06 | 3.1% |
| 28 | Other Pedestrian | -2.08 | 2.1% |
| 29 | Influenza & Pneumonia | -2.12 | 2.6% |
| 30 | Hot object/Substance | -2.21 | 0.2% |
| 31 | In situ and benign neoplasms, neoplasms of uncertain/unknown behavior | -2.31 | 1.2% |
| 32 | Other specified, classifiable Injury | -2.31 | 1.7% |
| 33 | Pneumonitis due to solids and liquids | -2.71 | 0.2% |
| 34 | Meningitis | -3.30 | 0.4% |
| 35 | Other transport | -3.33 | 0.1% |
| 36 | Certain conditions originating in the perinatal period | -3.39 | 1.3% |
| 37 | Motor vehicle traffic | -3.65 | 8.8% |
| 38 | Nephritis, nephrotic syndrome and nephrosis | -3.71 | 0.3% |
| 39 | Septicemia | -3.77 | 1.5% |
| 40 | Other specified, not elsewhere classified Injury | -3.80 | 0.7% |
| 41 | Fire/Flame | -6.12 | 2.9% |
| 42 | Fall | -7.01 | 0.4% |
| 43 | **Motor vehicle accident, as occupant<sup>^</sup>** | **-8.14** | **2.2%** |
| **All Deaths** |  | **-1.86** |  |

\*Average annual percent change
Total of 43 causes: 88.7%
Data source: CDC WONDER and CDC WISQARS<sup>4</sup>; causes with >50 deaths during 2002-2017
<sup>^</sup>ICD-10 Codes: V30-V39 (.4-.9), V40-V49 (.4-.9), V50-V59 (.4-.9), V60-V69 (.4-.9), V70-V79 (.4-.9), V83-V86 (.0-.3)

**Figure 4.**
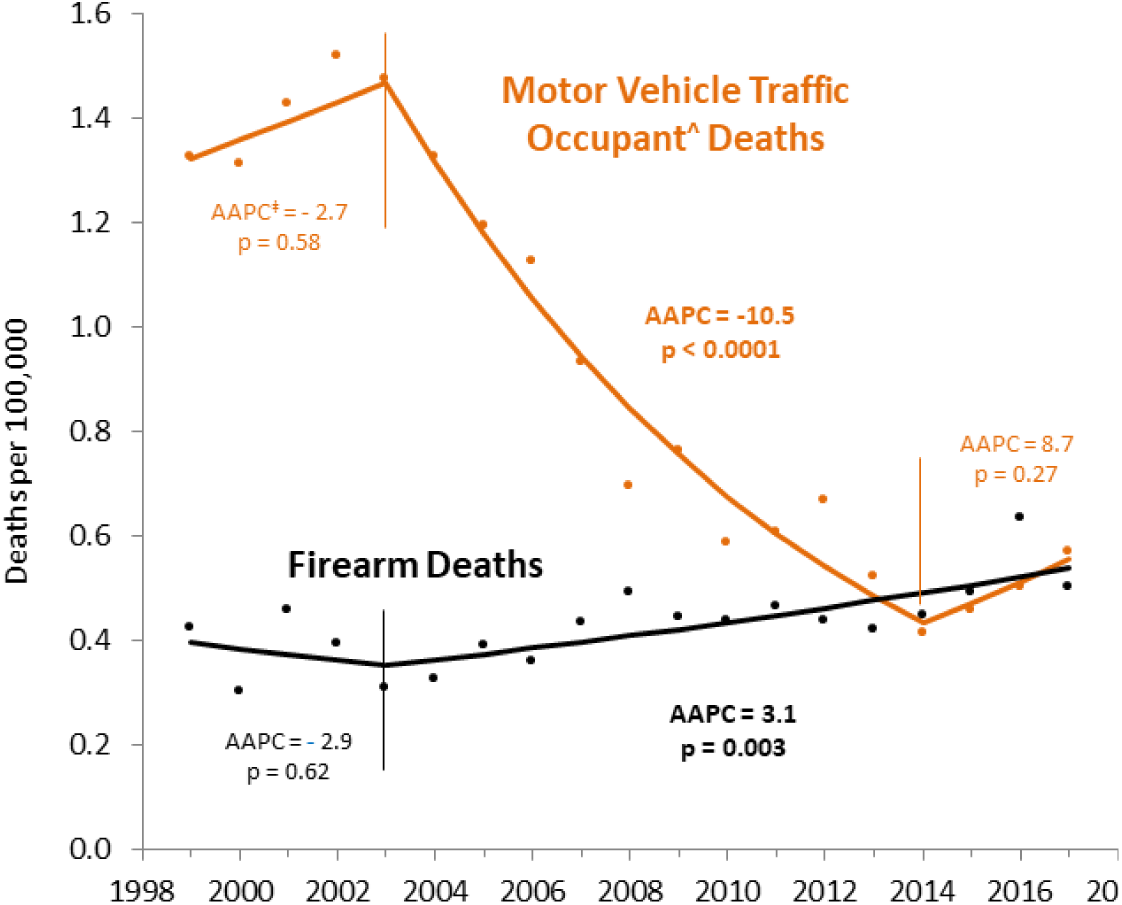
Joinpoint^9^/AAPC^‡^ Analysis of Annual Firearm and Motor Vehicle Traffic Occupant Death^^^ Rates, 1999-2017, U.S., Age 1-4. ^‡^ Average annual % change, based on constant variance (homoscedasticity) assumpt Data Source: CDC WISQARS^4^ **^^^** ICD-10 Codes:^8^ V3O-V39 (.4-.9),V40-V49 (.4-.9),V50-V59 (.4-.9), V60-V69 (.4-.9),V7O-V79 (.4-.9),V83-V86 (.0-.3)

### Correlation of Firearm Death Rate Trend in 1-4 Year-Olds with Firearm Sales

The national firearm background check rate also began to increase after 2004 and, thereafter, accelerated exponentially at an average AAPC of 8.7 during 2004-2017 (p < 0.0001) (Fig. 2**B**), concurrent with the increases in overall firearm death (Fig. 2) and fatal firearm death rates among 1-4 year-olds (Fig 2**A**). During those years, the firearm accident death rate in 1-4 year-olds and the annual firearm background check rate were directly correlated (R^2^ = 0.70, p = 0.0002) (Fig. 5**A**). The firearm accident death rate in 2017 for each legal jurisdiction (state and District of Columbia) was also correlated with its firearm background check rate for 2015-2017 (R^2^ = 0.17; p = 0.003) (Fig. 5**B**).

**Figure 5.**
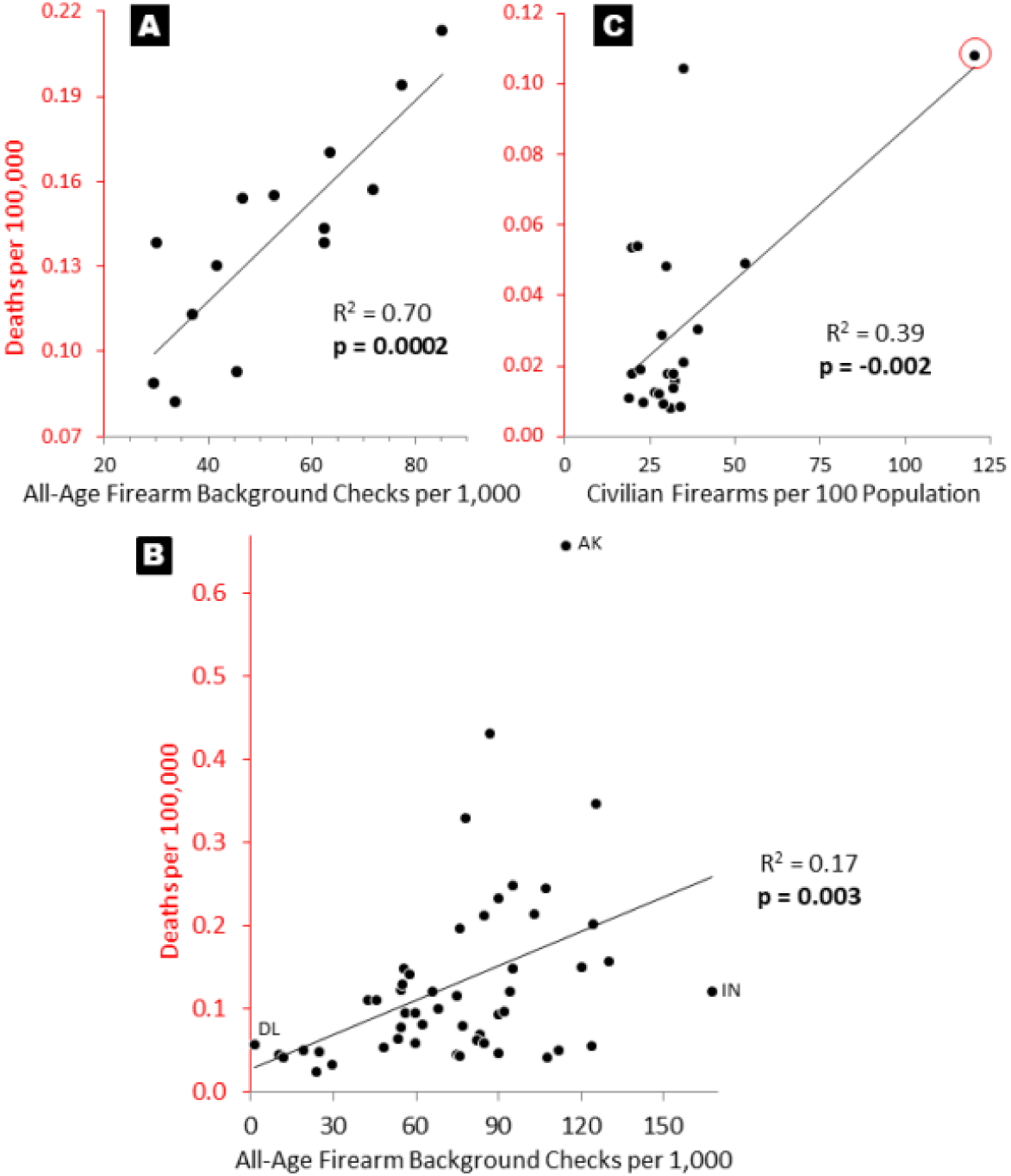
Correlation of 2017 Firearm Accidental Death Rate in 1-4 Year-Olds (y-axis) with: **A. All-Age Background Firearm Check Rate, Annually during 1999-2017, U.S.** <u>Data Source:</u> CDC WONDER,^4^ NICS^7^ **B. All-Age Background Firearm Check Rate, by U.S. State and D.C. during 2015-2017. AK-Alaska, IN-Indiana, DL-Delaware** <u>Data Source:</u> CDC WONDER,^4^ NICS^7^ **C. Estimated Civilian Firearm Prevalence in the 18 Countries with the Highest Prevalence during 2017 and Population >2 Million, excluding Iraq.** Red circle identifies U.S. <u>Data Source:</u> CDC WONDER,^4^ IHME GBD,^3^ Small Arms Survey^7^

### Type of Firearm Used in Deaths of 1-4 Year-Olds (Supplemental Fig. S1)

The available CDC WONDER data has handguns as the cause in 20%-30% of the fatal accidents, larger firearms (rifles, shotguns, assault weapons, etc.) in 5%–10% of the deaths, and 60%–80% not specified as to the type of firearm. The available data do not suggest changes in the proportion of types of firearm during 1999-2017. However, gun-related homicides in the age group do have a trend, with unspecified weapons increasing from 70%–80% during 1999-2004 to 90% by 2014.

**Supplemental Figure 1.**
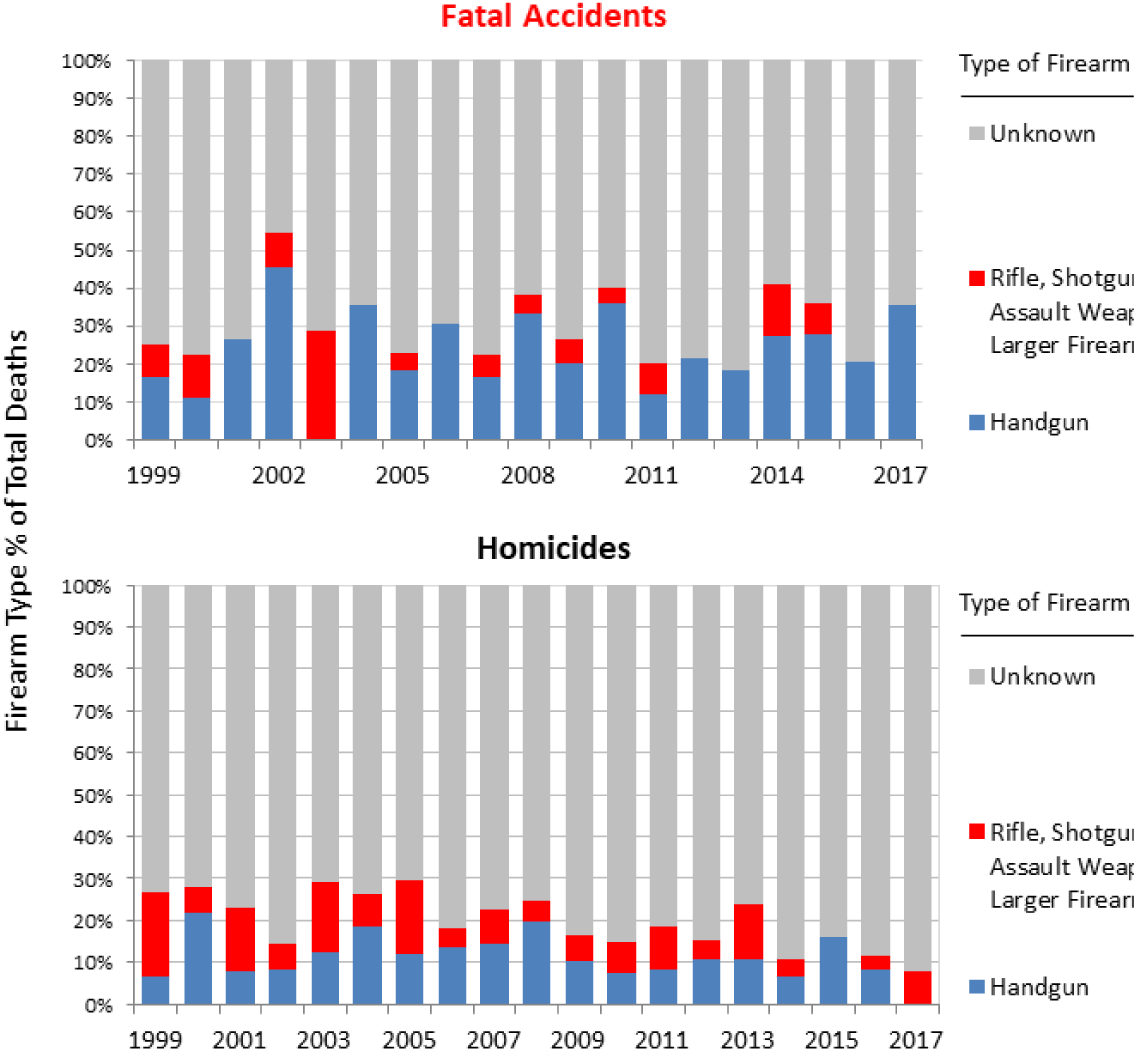
Annual Proportion of All Firearm Accidental Deaths (upper panel) and of All Homicides (lower panel) by Handgun, Larger Weapons* or by Unknown Type of Firearm, 1999-2017, Age 1-4. ^*^Rifle, shotgun, assault weapon, etc. <u>Data Source:</u> CDC WONDER^4^

### Firearm Death Rate Trend in 1-4 Year-Olds and the Federal Assault Weapon Ban

The U.S. had a Federal Assault Weapons Ban from 1994 until 2004 when it was not renewed because of equivocal evidence during the Ban that firearm deaths had decreased. In retrospect, firearm background checks decreased during the last 6 years of the Ban (Fig. 2**B**) and 1-4 year-olds had rate decreases during the Ban of fatal firearm accidents (AAPC = **−** 6.7, p < 0.0001), firearm homicides (AAPC = **−** 4.5, p = 0.02), and overall firearm deaths (AAPC = **−** 5.5, p = 0.001) (Supplemental Fig. S2). The year after the Ban expired and the national firearm background check rate began to exponentially increase, the rates of overall firearm deaths, firearm suicides, and fatal firearm accidents in 1-4 year-olds concurrently increased (Supplemental Fig. S2).

**Supplemental Figure 2.**
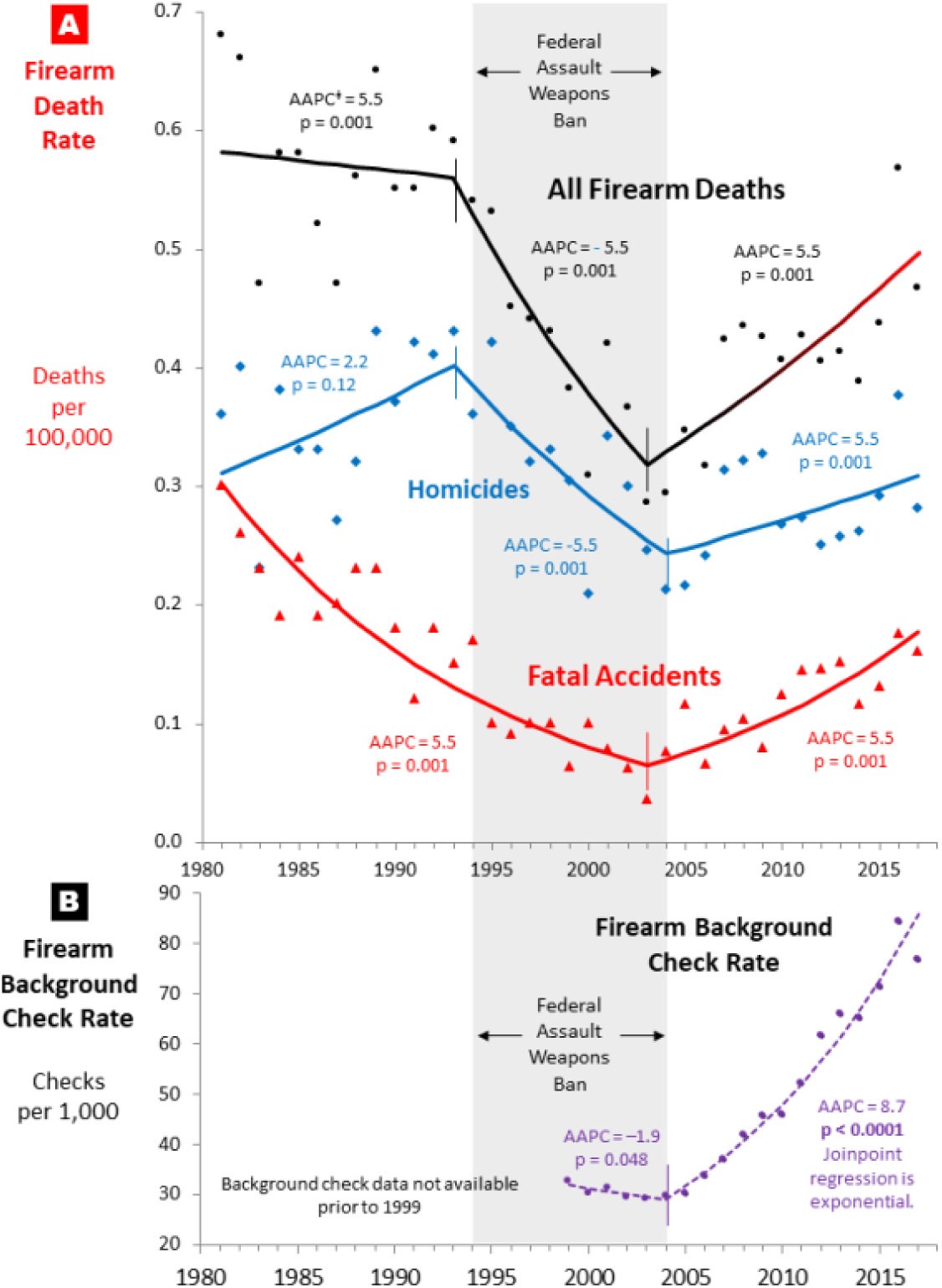
Joinpoint^9^/AAPC^‡^ Analysis of Annual Rates in U.S. of: A. Firearm Deaths In 1-14 Year-Olds, 1981-2017, by Type of Firearm Death B. All-Age Firearm Background Checks, 1999-2017. ^‡^ Average annual percent change Gray zone: Federal Assault Weapons Ban ^*^ based on constant variance (homoscedasticity) assumption <u>Data Source:</u> CDC WISQARS,^4^ NICS^6^

### Global Ranking of Firearm Accident Rate Trend in 1-4 Year-Olds

In terms of rate trends since 2002 when in the U.S. the firearm death rate in 1-4 year-olds began to increase (Fig. 2**A**), the U.S. had the greatest rate increase in the world during 2002-2017, according to IHME GBD data (Supplemental Table S1). Only 9 other countries had a consistent increase and each of the 189 countries in the rest of the world had a decrease (upplemental Table S1). The U.S. increase (AAPC = 6.01, p = 0.006) was greater than all of the countries with an increase, with the next greatest increase in Dominica (AAPC = 3.90). In terms of absolute rates during 2016-2017, the U.S. had the 2^nd^ highest firearm death rate (203/100,000) among 1-4 year-olds of 58 countries in Europe, North America, Australasia, and high-income Asia (Supplemental Table S2). Montenegro had the highest rate (217/100,000) and Greenland third (150/100,000). The 2017 national firearm accidental death rate in 1-4 year-olds was also directly correlated with the estimated civilian gun prevalence in the 18 countries with the highest rates of civilian gun ownership during 2017 and more than 2 million population (Fig. 5**C**) (p = 0.002).

## DISCUSSION

Our results indicate that an exponential increase in the firearm death rate of America’s youngest children explains in part why their overall mortality rate has ceased to improve like it has in other socio-demographically advantaged countries. Since 2014, 1- 4 year-olds in the U.S. were as likely to die of bullets as in motor vehicle accidents. Most of the firearm mortality increase in 1-4 year-olds was due to fatal firearm accidents that also increased exponentially during 2004-2017. If the trend continues, one in every100 deaths among 1-4 year-olds in the U.S. during 2019 will be due to firearm accidents. That both the overall and accidental firearm death rate increased while the overall and majority of causes of death decreased adds greater significance to the firearm problem.

Since 2004 when in the U.S. the Federal Assault Weapons Ban ended and firearm sales began to accelerate, the accidental firearm death rate in 1-4 year-olds was directly proportional to the increase in firearm background checks. The most recent absolute rates in firearm accidental deaths in the age group was highly significantly correlated geographically (at the state and D.C. level) with the concurrent corresponding all-age rate of firearm background checks. Also, the firearm accidental death rate in countries with the high civilian firearm prevalence was significantly correlated with the number of guns per civilian population. We therefore hypothesize that the increasing firearm death rates in young children that we obsevered was due to increasing firearm prevalence. Increasing violence displayed in video games and on television may have contrbuted to firearm death causation but only if firearms were accessible.

A critical limitation of our study is the ecologic limitation of attributing death rates to firearm sales rate in a cause-effect relationship. Although the correlations are not proof of causal relationship, the striking similarity of the joinpoint regressions of the overall, accidental, and undetermined firearm death rate profiles with the gun sales rate and the high degrees of correlation of accidental firearm death rates and firearm background check rates (0.0004 ≤ p ≤ 0.003) are highly suggestive of a causal relationship. Firearm accident mortality rates, including pediatric deaths, have previously been specifically linked to firearam sales and Google searches for buying and cleaning guns.^9^ Statistically significant or not, firearm prevalence and accidents is a logical synergism. There is also the assumption that firearm background checks are a surrogate for firearm sales. Background checks likely underrepresent the number of guns actually purchased since denials occur for <0.5% of checks, an approved buyer may purchase multiple firearms, and few states require checks for purchases from private parties. That the number of firearms in the home is not as predictive of suicide risk as the presence or absence of firearms^10^ also makes correlations more difficult to interpret to the extent that the increase in gun sales increased the number of guns per home more than the proportion of homes with guns. Also, the data available were not sufficient to analyze the role of other potentially important factors such as race and socioeconomic status.

Multilple comparisons have found states with more restrictive firearm legislation to have lower pediatric, unintentional, suicide, and overall firearm-related fatality rates, and more slowly rising suicide rates.^11–15^ A case-control study of firearms in events identified by medical examiner and coroner offices from 37 counties in Washington, Oregon, and Missouri, and 5 trauma centers in Seattle, Spokane, Tacoma, and Kansas City found that 4 practices of keeping a gun locked, unloaded, storing ammunition locked, and in a separate location are each associated with a protective effect for firarm injuries in homes with children and teenagers where guns are stored.^16^ Having a gun in the home is associated with an increased firearm death risk for everyone in the home, including <5 year-olds.^17^ General Social Survey gun ownership data indicates that handgun ownership in American families with young children increased to 72% in 2016 and is correlated with the increased death rate among young children from 2006 to 2016.^17^ Broadly reducing access to firearms has lowered firearm mortality in other countries.^18^

The Federal Assault Weapons Ban has been recently shown to have reduced mass shootings^19^ but not, to our knowledge, unintentional firearm deaths homicides in children. Assault weapons would not seem to be major cause of accidental deaths in young children. On the other hand, both handguns and long guns had progressively lower rates of permits issued during the last 6 years of the Ban (AAPC = –8.6, p = 0.03; AAPC = –4.8, p = 0.04, respectively) (Supplemental Fig. S3), suggesting that handguns were also affected by the Ban. Since the national firearm background check program began in November 1998, firearm sales may have been suppressed during the initial years of the ban, but the synchronicity of the Ban and sales also suggests that the Ban also suppressed sales of both handguns and long guns. Also, the low rate of handguns specified to have caused accidental deaths in 1-4 year-olds implies larger firearms were a primary method. In addition, the decreasing rate of handguns and increasing rate of unspecified weapons used in homicides of 1-4 year-olds suggests that the age group has been increasingly killed by larger firearms. The homicide trends are consistent with an increasing availability and use of larger weapons since the Federal Assault Weapons Ban expired in 2004. Thus the non-specified proportion of firearm accidental deaths may include an increasing proportion of larger weapons. Finally, with 16,000,000 1-4 year-olds in the U.S. and at least 16 million assault weapons (called *modern sporting rifles* by the National Shooting Sports Foundation^20^) in private ownership in the U.S.,^20^ the average of 1 assault weapon for every 1-4 year-old suggests that assault weapons by themselves could have contributed directly to the fatal firearm accidents in the age group. That the AR-15 (M16 style) rifle has become the most popular single model of rifle in the country^21^ adds to the feasibility of assault weapons accidentally killing the country’s youngest children.

**Supplemental Figure S3.**
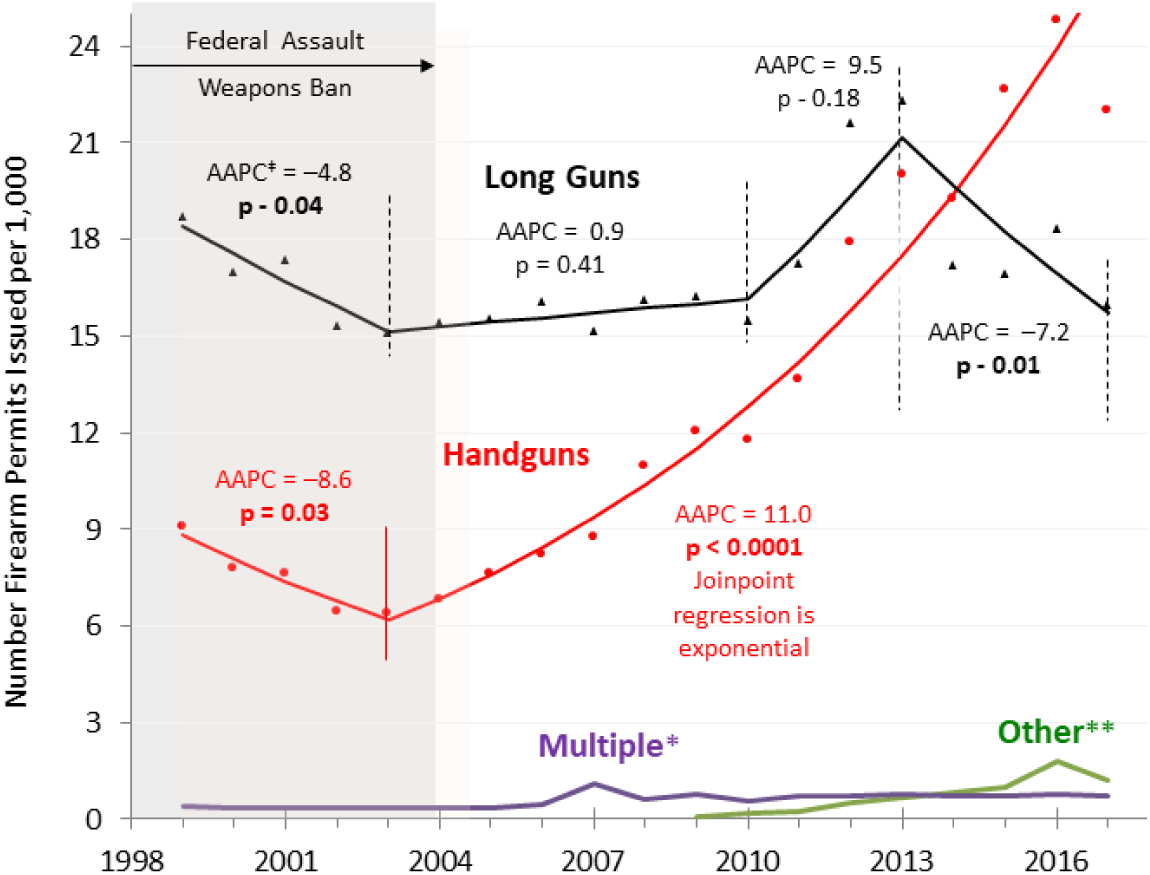
Joinpoint^9^/AAPC^‡^ Analysis of Annual Number of National Firearm Permits Issued, by Type of Firearm, 1999-2017. Gray zone: Last 6 year of Federal Assault Weapons Ban ^‡^ Average annual percent change, based on constant variance (homoscedasticity) assumption * Multiple (multiple types of firearms selected) ** Firearms that are neither handguns nor long guns (rifles nor shotguns) Data Source: N1CS^6^

The dramatic decrease in motor vehicle accident deaths, with the most recent data indicating that it is safer for 1-4 year-olds to be in cars than in homes with firearms, raises the issue of vehicle and firearm death comparisons. Federal funding of research was available to study vehicle deaths and prohibited for firearm research. Compared to requiring seat belt use, special safety seats for infants, airbag installation, power brake improvements and many other automobile safety features, firearms have had little such development. Requiring all drivers to be licensed and requiring motor vehicles to be parked in safe spots and their owners to be cited for parking violations has no parallel in the firearm world. These comparisons suggest what could be done to achieve a comparable reduction in firearm deaths.

The prominence of firearm deaths in American children requires addressing the problem of the country’s gun plethora.^22^ Of the world’s population, the 4% that live in America own nearly half of the entire global stock of 857 million civilian firearms.^23^ Eleven health professional organizations and the American Bar Association have recommended numerous national interventions that included protecting children from gunshot injuries.^24–28^ The American Academy of Pediatrics has conlcuded that “the absence of guns from homes and communities is the most effective measure to prevent suicide, homicide, and unintentional injuries to children and adolescents”.^29^ Because the gun lobby in the U.S. influenced legislation prohibiting government during the past 25 years from allocating resources to study the problem, the state of the science of firearm injuries among American children has lagged behind other areas of injury prevention.^30^ In 2017, the Firearm Safety Among Children and Teens (FACTS) Consortium, a National Institute for Child Health and Human Development-funded group of scientists and stakeholders, was formed to develop research resources for a pediatric-specific research agenda for firearm injury prevention.^31^

Ultimately, the non-fatal injury rate, which is multiple times higher than the death rate, must also be considered. In one study, for every person <18 years of age who died of bullets, 4 to 5 more were treated for gunshot wounds.^32^ The overall human and financial cost of firearm injuries and deaths in the U.S. is extraordinary.

## CONCLUSION

Young children in the U.S. have not had a decreasing mortality rate when compared to other high socio-demographic countries. A major reason is their rate of firearm deaths and especially fatal firearm accidents. Prior to 2004, the childhood firearm death rate did not increase during the Federal Assault Weapons Ban. Since then, the steadily increasing rate of sales and concomitant availability of, and access to, firearms in the U.S. has been associated with a dramatic increase in fatal firearm accidents in young children. The acceleration of firearm deaths and injuries among young Americans has become a national public health crisis that requires urgent and definitive solutions to address firearm prevalence.

Twelve health professional organizations and the American Bar Association have provided multiple recommendations, including funding of the CDC and National Institutes of Health to determine the most effective solutions. Ignoring these recommendation s and thereby allowing the increasing firearm death rate in young children to continue to increase is not a responsible option.

## Data Availability

All of the data are publicly available at websites of the U.S. Centers for Disease Control, Institute of Health Metrics and Evaluation of the University of Washington in Seattle, Washington, U.S., the U.S. Department of Justice, and the Small Arms Survey in Geneva, Switzerland, with specific sources to each provided in the References.

https://wonder.cdc.gov/ucd-icd10.html

https://webappa.cdc.gov/sasweb/ncipc/mortrate.html

ftp://ftp.cdc.gov/pub/Health_Statistics/NCHS/Publications/ICD10CM/2019/

https://www.fbi.gov/file-repository/nics_firearm_checks_-_month_year.pdf/view

http://www.smallarmssurvey.org/fileadmin/docs/T-Briefing-Papers/SAS-BP-Civilian-Firearms-Numbers.pdf

---

**Supplemental Table S1.**
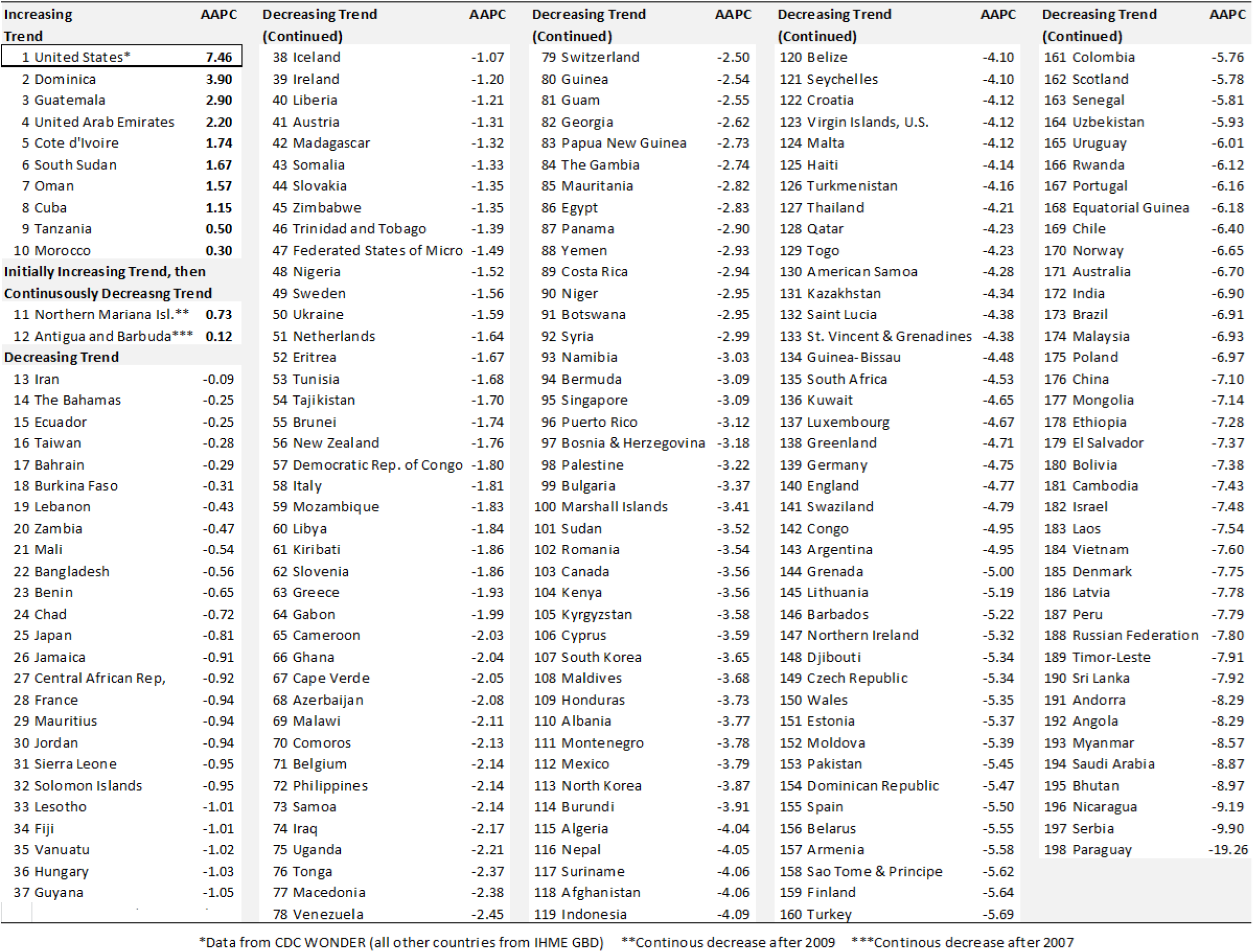
Global Ranking of Average Annual Percent Change (AAPC) in Unintentional Firearm Death Rate, 2002-2017, Age 1-4. <u>Data Source</u>: IHME GBD ^1^

**Supplemental Table S2.** Unintentional Firearm Death Rate, Age 1-4, 2016-2017, in North America, Europe, Australasia, High-Income Asia, by Rank Order. <u>Data Source</u>: IHME GBD^1^

|  | Country | Deaths<br>per<br>100,000 |  | Country | Deaths<br>per<br>100,000 |
| --- | --- | --- | --- | --- | --- |
| 1 | Montenegro | 0.217 | 30 | Israel | 0.018 |
| 2 | United States* | 0.203 | 31 | Germany | 0.018 |
| 3 | Greenland | 0.150 | 32 | Italy | 0.018 |
| 4 | Mexico | 0.066 | 33 | Poland | 0.017 |
| 5 | Bulgaria | 0.063 | 34 | India | 0.017 |
| 6 | Portugal | 0.057 | 35 | Finland | 0.016 |
| 7 | France | 0.055 | 36 | Denmark | 0.016 |
| 8 | Macedonia | 0.054 | 37 | Taiwan | 0.015 |
| 9 | Albania | 0.047 | 38 | Spain | 0.015 |
| 10 | Moldova | 0.045 | 39 | Japan | 0.015 |
| 11 | Slovakia | 0.045 | 40 | Iceland | 0.014 |
| 12 | Ukraine | 0.041 | 41 | Nepal | 0.014 |
| 13 | Romania | 0.038 | 42 | Northern Ireland | 0.014 |
| 14 | North Korea | 0.037 | 43 | Netherlands | 0.013 |
| 15 | Croatia | 0.031 | 44 | England | 0.013 |
| 16 | Serbia | 0.031 | 45 | Scotland | 0.012 |
| 17 | Malta | 0.030 | 46 | New Zealand | 0.012 |
| 18 | Andorra | 0.030 | 47 | Switzerland | 0.012 |
| 19 | Czech Republic | 0.027 | 48 | Wales | 0.012 |
| 20 | Latvia | 0.026 | 49 | Luxembourg | 0.011 |
| 21 | Belarus | 0.023 | 50 | Estonia | 0.011 |
| 22 | Greece | 0.023 | 51 | Hungary | 0.011 |
| 23 | Belgium | 0.022 | 52 | Australia | 0.010 |
| 24 | Brunei | 0.021 | 53 | Sweden | 0.010 |
| 25 | Canada | 0.021 | 54 | Slovenia | 0.009 |
| 26 | Lithuania | 0.021 | 55 | Norway | 0.009 |
| 27 | South Korea | 0.019 | 56 | Ireland | 0.009 |
| 28 | Austria | 0.019 | 57 | Cyprus | 0.008 |
| 29 | China | 0.018 | 58 | Bosnia and Herzegovina | 0.008 |
|  | Continued in next column |  | 59 | Singapore | 0.005 |

\*Data from CDC WONDER; Other countries from IHME GBD

